# Real Time Investigation of a large Nosocomial Influenza A Outbreak Informed by Genomic Epidemiology

**DOI:** 10.1101/2020.05.10.20096693

**Authors:** Waleed Javaid, Jordan Ehni, Ana S. Gonzalez-Reiche, Juan Manuel Carreño, Elena Hirsch, Jessica Tan, Zenab Khan, Divya Kriti, Thanh Ly, Bethany Kranitzky, Barbara Barnett, Freddy Cera, Lenny Prespa, Marie Moss, Randy A. Albrecht, Ala Mustafa, Ilka Herbison, Matthew M. Hernandez, Theodore R. Pak, Hala Alshammary, Robert Sebra, Melissa Smith, Florian Krammer, Melissa R. Gitman, Emilia Mia Sordillo, Viviana Simon, Harm van Bakel

## Abstract

**Background:** Nosocomial respiratory virus outbreaks represent serious public health challenges. Rapid and precise identification of cases and tracing of transmission chains is critical to end outbreaks and to inform prevention measures.

**Methods:** We combined conventional surveillance with Influenza A virus (IAV) genome sequencing to identify and contain a large IAV outbreak in a metropolitan healthcare system. A total of 381 individuals, including 91 inpatients and 290 health care workers (HCWs), were included in the investigation.

**Results:** During a 12-day period in early 2019, infection preventionists identified 89 HCWs and 18 inpatients as cases of influenza-like illness (ILI), using an amended definition, without the requirement for fever. Sequencing of IAV genomes from available nasopharyngeal (NP) specimens identified 66 individuals infected with a nearly identical strain of influenza A H1N1 (43 HCWs, 17 inpatients, and 6 with unspecified affiliation). All HCWs infected with the outbreak strain had received the seasonal influenza virus vaccination. Characterization of five representative outbreak viral isolates did not show antigenic drift. In conjunction with IAV genome sequencing, mining of electronic records pinpointed the origin of the outbreak as a single patient and a few interactions in the emergency department that occurred one day prior to the index ILI cluster.

**Conclusions:** We used precision surveillance to identify and control a large nosocomial IAV outbreak, mapping the source of the outbreak to a single patient rather than HCWs as initially assumed based on conventional epidemiology. These findings have important ramifications for more effective prevention strategies to curb nosocomial respiratory virus outbreaks.

## Introduction

Nosocomial outbreaks of pathogens represent major challenges for health care providers and institutions. It is critical for hospitals and health systems to not only quickly identify infected cases but also determine the source of the outbreak in order to mitigate the threat to patients and health care workers (HCWs). Nosocomial influenza virus outbreaks have been described worldwide (1-3); children, the elderly, institutionalized and immuno-compromised patients are particularly vulnerable. In some instances, nosocomial outbreaks have been caused by HCWs who work while ill (4).

Influenza virus is a single-stranded, negative sense, segmented RNA virus that causes an acute infection of the upper respiratory tract. Two main disease-causing influenza virus types, influenza A viruses (IAV) and influenza B viruses (IBV), circulate in human populations. IAV is further divided into subtypes based on the hemagglutinin (H) and neuraminidase (N) surface proteins. Both IAV subtypes H1N1 and H3N2 circulate in humans and were prevalent in the winter/spring of 2018-2019 in New York City (NYC).

Co-circulation and ongoing transmission of multiple virus subtypes in the surrounding community can complicate the determination of whether there is a clonal outbreak and make the identification of the source of the outbreak challenging. In order to develop effective strategies to control and prevent nosocomial outbreaks of influenza, approaches that enable rapid detection of transmission of a clonal influenza virus strain and identification of the origin of that transmission are needed. Complementing traditional epidemiological investigations, precision surveillance utilizing viral genome sequencing has been used previously to trace nosocomial influenza outbreaks (2, 5, 6).

Here we report the integration of our conventional infection prevention measures with precision surveillance, including sequencing the genome of IAV from clinical biospecimens and data mining of electronic medical records (EMRs), to successfully identify and control a large nosocomial IAV outbreak affecting both inpatients and HCWs.

## Results

### Epidemiology of the nosocomial influenza outbreak

In early 2019, symptoms suggestive of influenza like illness (ILI) were first observed in several HCWs as well as in inpatients receiving critical care at Hospital A in NYC. At the direction of the Infection Prevention Department, the hospital’s incident command system was activated. An extensive outbreak investigation was started, which included mandatory staff symptom checks and testing of all inpatients with any respiratory symptoms, regardless of fever status. Enhanced cleaning of patient care areas and clinical staff workspaces was also performed.

Over the course of the hospital-wide outbreak investigation, a total of 381 individuals (91 inpatients and 290 HCWs) were screened by regular body temperature checks, symptom surveys and/or molecular diagnostic testing for IAV, IBV and respiratory syncytial virus (RSV). A total of 18 inpatients (19.8%) and 89 HCWs (29.7%) included in the epidemiological investigation tested positive for IAV (**Figure 1A**).

**Fig 1:**
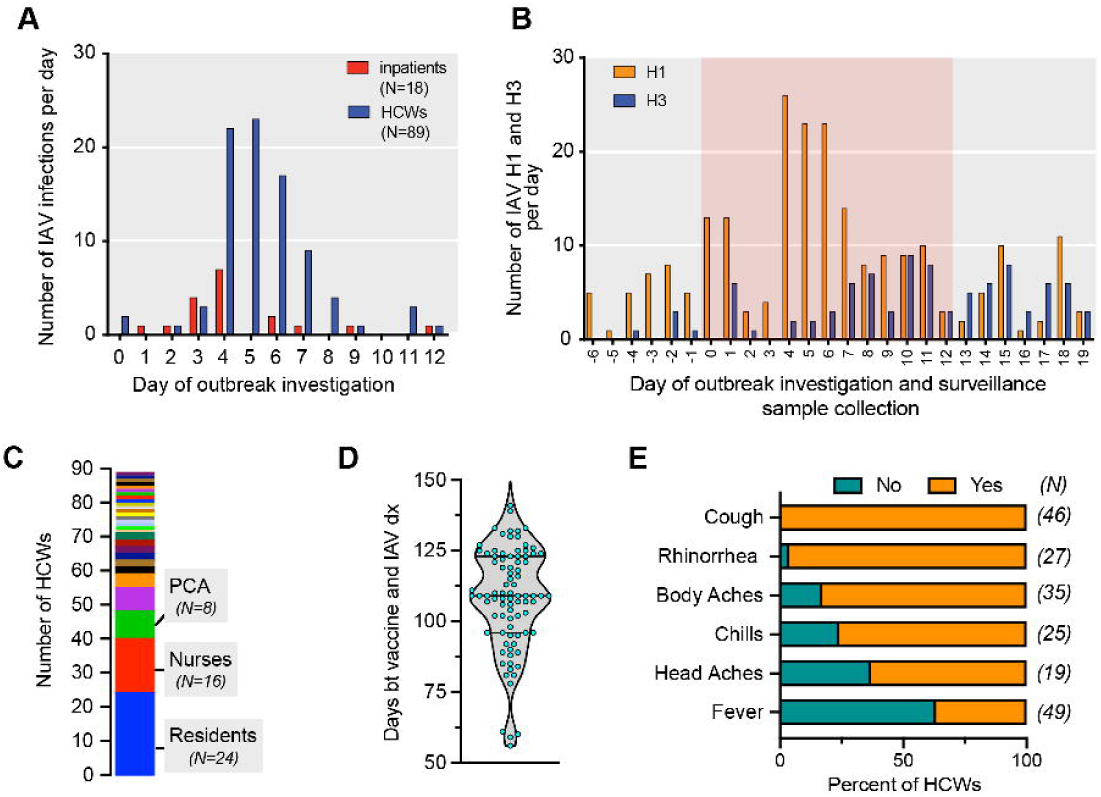
Epidemiology of the nosocomial IAV outbreak. **A:** Time line of the nosocomial IAV outbreak at a metropolitan hospital. Day 0: initiation of the infection prevention investigation. IAV, influenza A virus; HCW, healthcare workers. **B**: Distribution of IAV subtypes detected in individuals identified in the outbreak investigation and patients seeking care at the hospital and testing positive for IAV between 6 days before (Day -6) and 19 days after (Day 19) the initiation (Day 0) of the infection prevention investigation. The red background highlights the period of the outbreak investigation. **C**: The distribution of professions (29 job categories) of the 89 healthcare workers who tested IAV positive. PCA: Patient care assistant. **D**: The distribution of days between receiving seasonal influenza virus vaccination and being tested positive for IAV among HCWs. Of note, 87 of 89 HCWs tested positive for IAV were vaccinated. **E**: Clinical signs and symptoms reported by HCWs who were tested positive for IAV. The data available for each symptom differs with respect to the number of employees (N listed in parenthesis provides the absolute numbers). Note that 63% of HCWs were afebrile.

Subtyping of IAV from the nasopharyngeal (NP) samples collected during the epidemiological investigation (N=104), the routine influenza surveillance at Hospital A (N=150) and Hospital B (N=231), revealed a stark increase of IAV/H1 at day 4, 5 and 6 of the investigation (**Figure 1B**). Of note, all the samples from inpatients and HCWs included in the investigation that we successfully subtyped harbored IAV/H1N1, suggesting a single transmission chain.

The 89 positive HCWs were distributed across 29 different work assignment categories (**Figure 1C**), predominantly front line care providers, including 24 resident physicians (residents, fellows, or interns), 16 registered nurses, 8 patient care assistants, and 6 attending physicians. Eighty-seven of these 89 HCWs (>90%) had been vaccinated with the quatrivalent seasonal influenza virus vaccine two to five months (average: 108 days) prior to being tested positive for IAV (**Figure 1D**). Importantly, these infected HCWs presented various, and mostly minor, clinical symptoms, and most individuals would not have been classified as having "influenza-like illness” given the lack of fever (**Figure 1E**). Because of this altered influenza disease manifestation in vaccinated HCWs, the case definition was amended early in the context of our investigation.

### Genomics of the nosocomial influenza virus outbreak

In order to determine whether there was transmission of a single IAV strain or there were several independent introductions into the hospital system, we performed next generation sequencing (NGS) of IAV from the NP specimens that were banked following the initial diagnostic testing. As part of our Institution’s Pathogen Surveillance Program, we routinely sequence influenza virus from a subset of the patients seeking care at our hospitals (termed “surveillance”). Thus, in addition to cases identified in the outbreak investigation, we included surveillance samples obtained from the general patient population seeking care at our hospitals as part of our Institution’s Pathogen Surveillance Program in order to determine potential community circulating strains.

Complete genomic sequences were obtained from 214 IAV isolates (**Figure 2A**), including 126 from Hospital A (investigation and surveillance) and 88 from Hospital B (surveillance only). Pairwise comparison of these viral genomic sequences showed a large cluster of 66 viral isolates that differed by no more than 3 single-nucleotide variants (SNVs), indicating that a single virus clone was responsible for a large portion of the nosocomial outbreak (**Figure 2B**). Additionally, our analyses indicated that other independent introductions of IAV H1N1 strains, with limited forward transmissions, had caused smaller clusters of ILI at both Hospital A and Hospital B. We also noted two small independent clusters due to transmission of IAV H3N2 viruses.

**Fig 2:**
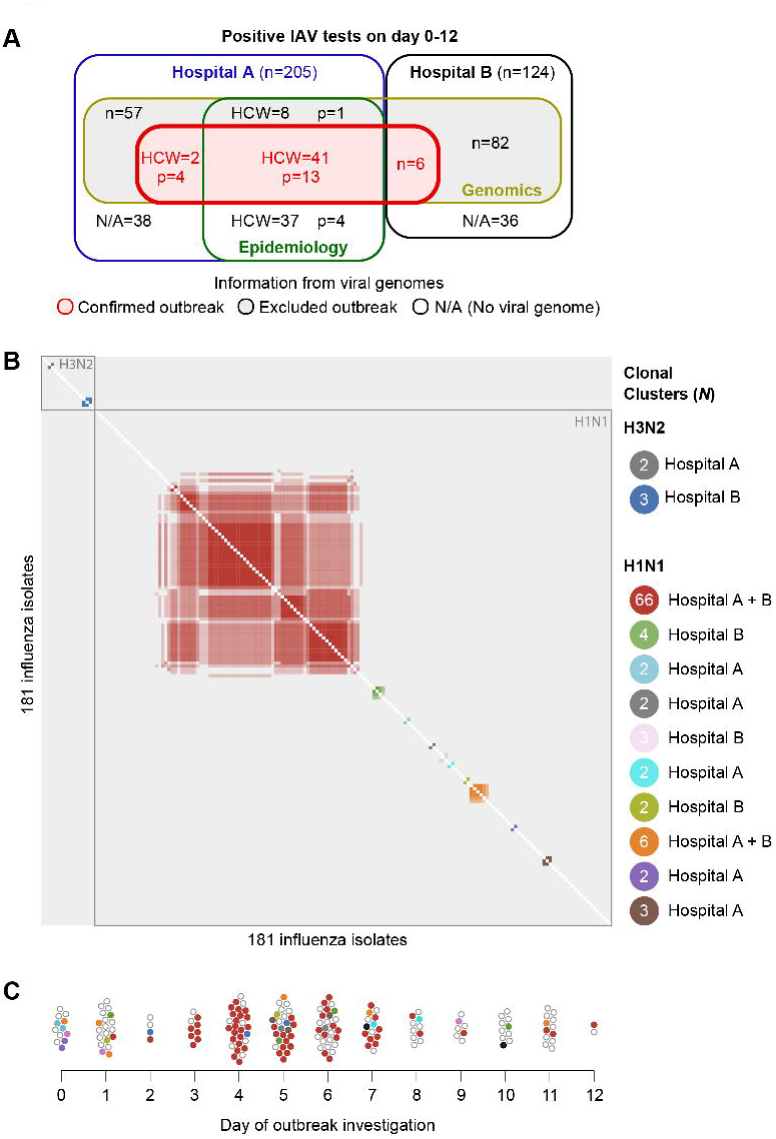
Genomics of the nosocomial outbreak. **A:** Venn diagram illustrating the number of sequenced outbreak confirmed H1N1 strains (N=66) and outbreak excluded IAV, including the unrelated H1N1 strains (N=113) and H3N2 strains (N= 36), identified in Hospital A (investigation and surveillance) and Hospital B (surveillance). Epidemiology describes the cases identified by infection prevention. **B**: Pairwise comparison of the complete viral genomes. Note the tight cluster of the outbreak H1N1 strains (N=66, red cluster) at the center and the two small H3N2 clusters at the top left of the pairwise comparison. **C**: Dynamics of the case numbers and IAV strains during the investigation period. All color codes used in this panel are same as those used in the panel B. The outbreak H1N1 strain (shown in red) was first detected on the day after the initiation of the investigation in a hospitalized patient. The first two employees who were tested positive for IAV on the day of the initiation of the investigation harbored unrelated H1N1 strains.

Correlating virus genomic sequences with the timing and the source of these isolates showed that all of the virus isolates obtained on Day 0 and most of virus isolates on Day 1 of the infection prevention investigation were distinct from the viral isolate that caused the large outbreak. Although two HCWs were tested positive for IAV on Day 0, their viruses were different from the one that caused the outbreak, and not associated with any nosocomial transmission. All HCWs infected with the outbreak virus had received the seasonal influenza virus vaccine. The first isolate that clustered with the outbreak virus strain was obtained from a patient identified on Day 1 of the investigation (**Figure 2C**).

Altogether, the genomic analyses of available clinical influenza isolates showed that cases identified by the conventional epidemiological investigation encompassed patients and HCWs who together harbored 12 different IAV strains and that only one specific strain of these 12 caused widespread nosocomial transmission.

### Phylogenetic and functional properties of the IAV outbreak strains

Phylogenetic analysis of the sequenced IAV genomes showed that the outbreak strain tightly clustered within a specific H1N1 6b1A subclade. Other IAV H1/N1 isolates obtained in the infection prevention investigation and routine surveillance were mapped throughout the H1N1 6b1A clade (**Figure 3A**, compare green to red dots) and likely reflected the predominant seasonal spread in the community. A detailed analysis of genomic sequences of all outbreak IAV strains showed that they were highly conserved; most of nucleotide variations occurred in the HA and the NA segments encoding surface proteins and the majority of predicted amino acid changes occurred only in the NA segment (**Figure 3B**). We selected five outbreak virus strains with representative variants in to be propagated in cell culture for functional characterization (**Figure 3C**). Hemagglutination inhibition assays performed with these outbreak virus strains confirmed that none had drifted as compared to the H1N1 vaccine strain used in that season (**Figure 3D**).

**Fig 3:**
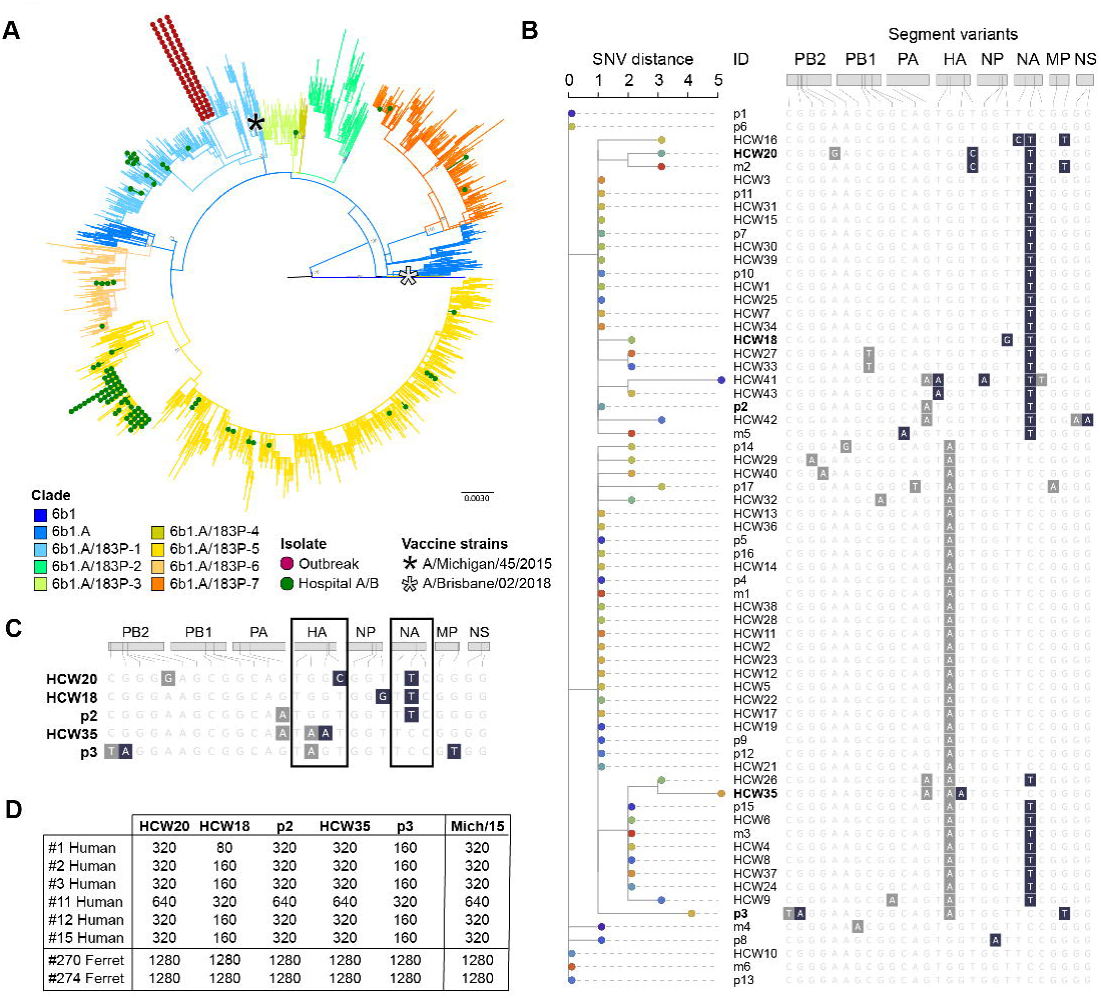
Phylogenetic and functional properties of the IAV outbreak strains. **A:** Analysis of IAV H1N1 genomic sequence diversity. Note that the outbreak strains form a tight cluster that maps to one subclade of the H1N1 6b1A clade whereas those not responsible for the outbreak are diverse and map to various subclades that are different from the outbreak one. IAV H1N1 vaccine strains A/Michigan/2015 and A/Brisbane/2018 are included in the analysis as references. **B**: Phylogenetic relationships based on single nucleotide variant (SNV) distance among the outbreak virus strains. All eight viral segments are shown, with grey indicating synonymous changes and dark blue indicating non-synonymous substitutions. **C**: Genotype of the five representative outbreak strains that were grown in cell culture. Grey, synonymous mutations; dark blue: non-synonymous changes. **D**: Hemagglutination inhibition titers of five representative outbreak strains with the sera of six recently vaccinated individuals. Sera from ferrets infected with the vaccine strain A/Michigan/45/2015 served as a positive control for antisera and the A/Michigan/45/2015 virus is the antigen positive control.

### Reconstruction of the transmission chain in the early days of the outbreak

In order to understand the origin of the outbreak and the factors that facilitated its rapid spread, we first focused on the early stages of the outbreak between days 0-3. We obtained IAV genomes for 10 of the 12 positive cases identified in the outbreak investigation (83%), as well as for 34 of 46 other positive surveillance samples obtained at hospitals A and B (74%). Of these, eight cases and one surveillance sample matched the outbreak strain, and we used the *PathoSPOT* framework (https://pathospot.org) to query various electronic hospital records in order to create a timeline (**Figure 4A**).

**Fig 4:**
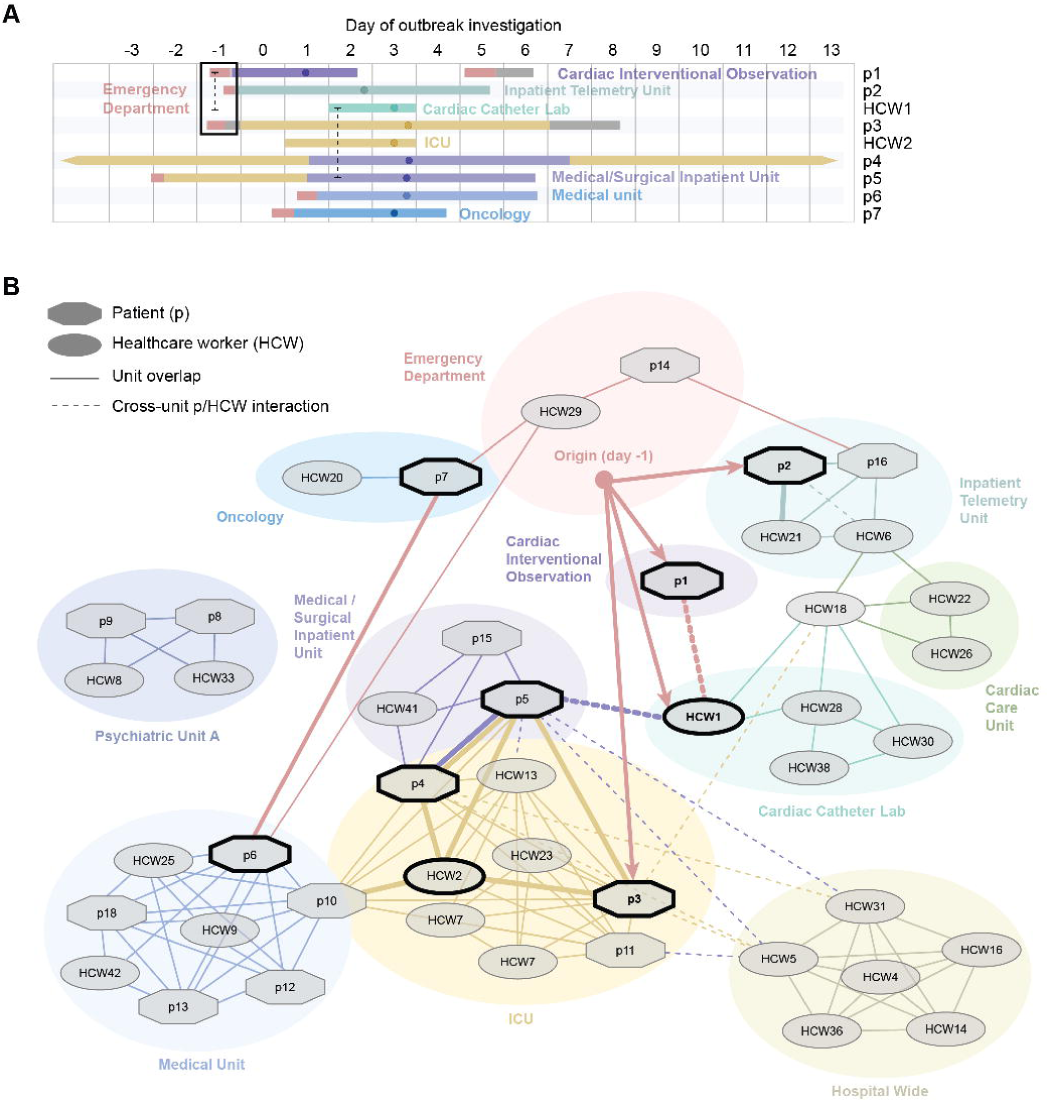
Reconstruction of the transmission chain in the early days of the outbreak. **A:** Timeline and locations inside the hospital of the first nine cases harboring the IAV/H1N1 outbreak strain. Day 0 is the day the investigation started. Dots on the line point to the day on which patient/health care worker tested positive for IAV. The dashed lines indicate presumed contact between infected individuals. The black box highlights the three patients that received care in the emergency department a day prior to the start of the infection control investigation. **B**: Interaction network of health care workers (HCW) and patients (p) who were tested positive for the IAV H1N1 outbreak strain. The cases identified by thick black outlines were critical in seeding the transmission within the hospital.

The data showed that 4 of the 9 initial nosocomial IAV cases were seen (patients p1, p2, and p3) or worked (HCW1) in the emergency department (ED) on the same day (Day -1), during overlapping time periods. The 3 patients were admitted from the ED to different wards and had no other shared interactions with HCWs, indicating that the common exposure most likely occurred in the ED. Similarly, because one of the patients who acquired nosocomial IAV did not have direct contact with HCW1 and had already been transferred out prior to the time HCW1 was present in the ED, the evidence indicates HCW1 was exposed during that work shift rather than being the primary case.

In contrast, patient p3, the putative primary case for this outbreak, was brought to the ED in the morning of Day -1, several hours before p1 and p2, and interacted with HCW1. P3 was admitted to a medical unit the same day (**Figure 4A**, grey) with fever, shaking chills, dyspnea, and abdominal pain, but developed systemic inflammatory response syndrome and was transferred to the ICU where the patient was intubated, a procedure that can generate significant aerosols (7) Because blood cultures of p3 grew Gram-negative bacteria, the diagnosis of IAV was delayed. However, the patient remained febrile despite antibiotics prompting diagnostic testing for influenza virus on day 3.

The next three early cases (HCW2, p4 and p5) most likely acquired infection in the ICU from p3, although p4 and p5 had further overlapping stays following transfer to the same medical/surgical inpatient unit from the ICU. Our data suggests that cases p6 and p7, who were admitted through the ED several days after the start of the outbreak, acquired IAV infection from ED HCWs who had been exposed to p3 and became IAV-test positive in the days thereafter.

The interaction network based on available contact records (**Figure 4B**) indicates that almost all later cases can be traced back in the some way to the initial nine cases shown in the timeline (**Figure 4A**), with transfers of patients acting as the major vectors for spread to other hospital units. A direct transmission link could not be documented for two patients cared at and two HCWs assigned to the same closed unit psychiatric ward, suggesting that indirect interactions may have occurred elsewhere in the hospital.

## Discussion

In this report we describe using a combination of conventional infection prevention approaches and precision surveillance to identify and control a nosocomial IAV outbreak in the spring of 2019. Swift interventions such as widespread molecular influenza virus testing and modification of the case definition to include mild respiratory disease presentations brought the outbreak under control within two weeks.

Almost a third of the 290 HCWs included in the epidemiological investigation tested positive for IAV (**Figure 1**). The amended case definition for ILI was based on the realization that most HCWs infected with IAV were afebrile. The vast majority of infected HCWs had received the seasonal influenza virus vaccination. Among the infected hospital employees for which viral influenza genotypes were available (49/89), 41 harbored the outbreak strain whereas eight were infected with unrelated H1N1 viruses (**Figure 2**). In the absence of our infection prevention intervention, many of these cases would have gone undiagnosed, pointing to the fact that influenza virus infections in vaccinated HCWs remain largely under-diagnosed due to the milder disease presentation. Thus, diagnostic testing of vaccinated HCWs with mild symptoms should be considered, especially when the seasonal influenza vaccine is well matched to the circulating viruses.

By sequencing influenza virus genomes from the infected patients and HCWs, we could focus the investigation into the source of the outbreak on only those cases that actually were infected with the identified outbreak virus strain. Of note, the two HCWs who were the first to be diagnosed with ILI and initially considered the likely source(s) of the outbreak were infected with viruses distinct from the outbreak strain (**Figure 2**). It is also important to note that the time of diagnosis may not be a good indicator for the actual chronology of an outbreak, which may be true especially for vaccinated HCWs who may ignore or downplay symptoms. Indeed, we observed that almost three days transpired between the onset of symptoms and positive IAV test in HCWs compared to the average of one-half day for inpatients. Integrating data from various hospital electronic records with molecular confirmation of which patients and HCWs were infected with the outbreak strain enabled reconstruction of the dynamics of the outbreak, identification of the likely primary case, and therefore ensured reassessment of transmission and heightened remediation for areas where transmission occurred.

Since we routinely sequence influenza virus isolates from patients receiving care at our health system as part of our Pathogen Surveillance Program, we could compare the strains from the outbreak investigation conducted at Hospital A to the strains found in the surveillance of Hospital A as well as Hospital B (**Figure 3**). These additional data allowed us to not only identify previously unrecognized smaller transmission events (four inpatients and two HCWs at Hospital A and six patients at Hospital B) but also ascertain that there was a large (in number) but limited (in time) outbreak of H1N1 in our health system (**Figures 2 and 3**). Importantly, this specific H1N1 virus outbreak strain did not spread further in the community.

A limitation of our study is that we did not have access to biospecimens from 22 HCWs whose tests were performed at laboratories outside our health system. Additionally, only partial viral genomes could be retrieved from two of the available biospecimens linked to the epidemiological outbreak investigation. However, we were able to obtain viral genomes for all the patients identified during the first three days of the outbreak, providing a solid foundation for the reconstruction of the transmission chain.

Our data suggest that the outbreak began in the ED most likely through introduction of the virus by a single patient, who had received aggressive resuscitative care and was subsequently transferred to the intensive care unit of Hospital A (**Figure 4)**. A possible solution to mitigate such risks to HCWs and patients in the future is to enhance screening and isolation of patients coming into the ED with any respiratory symptoms, even when an alternative diagnosis seems to be the predominant complaint. Additionally, recognition by hospital leadership of the potential for transmission even from HCWs with mild influenza illness resulted in administrative support for intensified education of staff to avoid working while ill, extended sick leave when needed, and a move away from the HCW culture of “presenteeism” which can contribute to nosocomial transmission of influenza (4) and other respiratory viruses.

It is critical for patient care that any healthcare organization quickly detects the occurrence of hospital acquired infections and limits their spread through swift identification of their origins. Conventional infection prevention approaches, however, are challenged if the hospital outbreak occurs in the context of widespread community acquired infections (e.g., during the peak of the influenza season as in this study). Our findings are applicable to a wide range of highly transmissible respiratory viral pathogens including Severe Acute Respiratory Syndrome Coronavirus 2 (SARS-CoV-2). Indeed, emerging evidence suggest that a high percentage of HCWs became infected with SARS-CoV-2 before screening for acute infection became more available. Implementation of precision surveillance measures as outlined here will be of critical importance to mitigate the risks of nosocomial transmission for patients and HCWs alike.

## Methods

### Ethics statement

The Pathogen Surveillance Program and the in depth analysis of the outbreak were approved by the Institutional Review Board of the Icahn School of Medicine at Mount Sinai.

Summary of infection prevention measures and investigation.

When we detected ILI cases in inpatients in early 2019, infection prevention measures, including extensive screening of inpatients and HCWs for ILI and collection of nasopharyngeal (NP) swab specimens were implemented. Once the outbreak was identified, a case definition was developed. This specific case definition included any HCWs or patient with cough, rhinorrhoea, sore throat, body aches, with or without fever, and positive diagnostic test for influenza virus by molecular polymerase chain reaction (PCR) testing (Xpert® Xpress Flu/RSV test, Cepheid) of a NP swab specimen in universal transport medium (NP-UTM).

During this initial investigation, a total of 381 individuals from hospital A, including 91 inpatients and 290 HCWs, were screened based on symptoms and laboratory detection of respiratory viruses. Among these 381 individuals, a total of 89 HCWs and 18 patients were found to be infected with IAV. A high incidence of cases was found among HCWs, including interns, residents, fellows, and rotators, as well as nursing and associated fields. Clusters were found in several areas inside the hospital including intensive care units and several medical/surgical floors and an inpatient psychiatric unit. Prophylaxis with oseltamivir was offered to exposed HCWs and inpatients. Treatment with oseltamivir was provided to infected inpatients and HCWs, in addition to extended sick leave for HCWs who remained afebrile but symptomatic. There was no mortality reported in association with the outbreak. One patient required readmission secondary to ILI. We educated staff throughout the hospital including the trainees on symptoms associate with influenza and encouraged all to report to their supervisors if they had any symptoms. Mandatory symptom checks at beginning of each shift were also implemented. Anyone symptomatic was encouraged to get tested. We were able to contain this outbreak from detection to eradication within 10 days, with incredible collaboration between many groups.

### Collection of NP-UTM for influenza A subtyping and sequencing

A total of 486 IAV positive NP-UTM specimens, including 104 samples from the epidemiological outbreak investigation, 150 routine influenza surveillance samples at Hospital A, and 231 routine influenza surveillance samples at Hospital B, were collected and submitted for IAV subtyping and next generation sequencing (NGS). The time frame from which surveillance and investigation samples were included covered a total of 27 days, starting from six days before the investigation to seven days after the 12 days long outbreak investigation. Viral RNA was extracted from 280μL of NP-UTM using the QIAamp Viral RNA Minikit (QIAGEN, cat. 52904), as per the manufacturer’s instruction. To distinguish between IAV H1 and H3, we used a modified version of the WHO One-Step Real-time RT-PCR protocol and viral RNA isolated from NP-UTM specimens. The primers and probes are multiplexed so that one can distinguish influenza A subtypes (e.g., IAV/H1pdm09 and IAV/H3) in the same reaction. All reactions were run in duplicates using the QuantiFast Pathogen RT-PCR +IC Kit (QIAGEN, cat. 211454) on the Roche LightCycler 480 Instrument II (Roche Molecular Systems, 05015243001) in 384-well plates. Every run included two positive controls and nuclease-free water served as a non-template control. The following temperature profiles was used: 50°C for 20 min, 95°C for 1 sec, 95°C for 5 min, followed by 40 cycles of 95°C for 15 sec and 60°C for 45 sec, during which quantitation of products occurred. For a result to be called positive for influenza A(H1)pdm09 or A(H3) subtype, replicates were required to have Ct values < 35. A result was deemed negative if both reactions displayed Ct values ≥ 35. A result was deemed inconclusive if one of the reactions was negative (Ct ≥ 35).

### Next generation sequencing of the genome of influenza virus

RNA from the NP samples and viral isolates were used for whole genome amplification of the IAVs by performing a multisegment RT-PCR for whole genome amplification with Opti 1 primers (8). Multi-segment PCR amplicons were cleaned by 0.45X of Agencourt AMPure XP magnetic beads (Beckman Coulter) according to manufacturer’s protocol. The concentration of purified amplicons was measured using the Qubit High Sensitivity dsDNA kit. After samples were normalized to a concentration of 0.2 ng/μl, adapters added by tagmentation using the Nextera XT DNA library preparation kit (Illumina). Samples were purified using 0.7X of Agencourt AMPure XP Magnetic Beads and fragment size distributions were analyzed on a Bioanalyzer using the High Sensitivity DNA kit (Agilent). After bead-based normalization (Illumina) according to the manufacturer’s protocols, sequence-ready libraries were sequenced in a paired-end run using the MiSeq v2, 300cycle reagent kit (Illumina). Complete genomes were assembled from next generation Illumina reads using a custom genome assembly pipeline (8). Custom codes and pipelines are also available at https://bitbucket.org/bakellab/flugap.

### Identification of clonal outbreak isolates

To detect clusters of highly related outbreak isolates we used the open-source *PathoSPOT* (<u>Patho</u>gen <u>S</u>equencing <u>P</u>hylogenomic <u>O</u>utbreak <u>T</u>oolkit) software (https://pathospot.org), which we developed to aid detection and visualization of transmissions in nosocomial settings. The *PathoSPOT compare* pipeline was used to first cluster influenza genomes based on their Mash distance using default distance threshold parameters. Segment sequences were concatenated together into a single genome sequence per isolate, and the concatenated sequences were then used to construct a multi-genome alignment of isolates in each cluster using *parsnp*. (9) Pairwise distances between genomes were then calculated as the number of single nucleotide variants (SNVs) between genome alignments in each cluster for further analysis. To identify transmission events we used the *PathoSPOT heatmap view* (**Fig. 2**). We set a threshold of ≤3 SNVs to identify potential transmissions. Next, the *dendro-timeline view* was used to perform phylogenetic analysis of outbreak isolates, and to reconstruct the early outbreak timeline based on the full admission/transfer/discharge (ADT) history for each patient obtained from the electronic medical record. Finally, the ADT history was combined with patient-HCW interaction data to reconstruct a network of all known contacts in *Cytoscape*. (10).

### Phylogenetic analysis

Hemagglutinin (HA) H1 segment sequences representing the global diversity of H1N1 viruses circulating during the 2018-19 influenza season (September 1, 2018 - April 30, 2019) as of August 20, 2019 were obtained from the Global Initiative on Sharing All Influenza Data (GISAID) database. Only complete HA coding sequences (1701 nt) were included. In order to avoid potential duplicates, for records with identical strain names but different accession numbers, only one sequence was retained. When available, sequences derived from the original samples or from isolates with the lowest number of passages were included. After applying these filters, a total of 8,314 HA H1 sequences were retained. In addition, to inform lineage annotation, 6 reference strains from 2015 to 2018, including the vaccine strains A/Michigan/45/2015 (H1N1) and A/Brisbane/02/2018 (H1N1) were included. To enable comparison, 163 H1N1 sequences from isolates recovered from Hospital B patients and HCWs during the outbreak time-window were also included. A maximum-likelihood (ML) phylogeny was inferred with RAxML (11) performing 20 ML searches under the GTRCAT model of nucleotide substitution and assessed with 100 bootstrap replicates.

In order to confirm phylogenetic grouping, we inferred a second ML analysis at the whole genome level for all strains that belonged to the highest supported clade that contained the outbreak isolates. For this analysis, the coding regions for the 10 main IAV proteins (PB2, PB1, PA, HA, NP, NA, M1, M2, NS1, and NEP) were concatenated. In addition to the outbreak isolates, this clade included 17 isolates from locations outside New York City that were retained as an outgroup.

### Propagation of clinical viral isolates

NP-UTM that tested positive for influenza virus were aliquoted and frozen at -80°C. For viral isolation and growth, samples were pre-diluted in infection media consisting on Minimum Essential Media (Gibco) supplemented with 1μg/ml of tosyl sulfonyl phenylalanyl chloromethyl ketone (TPCK) treated-trypsin (Gibco) and added to 90% confluent MDCK cells monolayer. Infections were let to proceed for 1 hour with intermittent shaking. The inoculum was removed and infection media was added. Plates were incubated at 37°C with 5% CO_2_ and supernatants were collected at 24-48 hours after infection when cytopathic effect was observed. Samples were cleared by centrifugation at 4300g for 5 min, supernatants were filtered to avoid bacterial contamination then stored at -80**^o^**C.

### Hemagglutination inhibition (HI) assay

These assays were performed as described before (12, 13). Briefly, serum samples from eight healthy study participants receiving the seasonal influenza virus vaccine (four weeks post-vaccination) were treated with receptor destroying enzyme (RDE) (14) and serially diluted (2 fold dilutions) in 96-V-well microtiter plates, followed by addition of 8 hemagglutination units (HAU) from five selected viral isolates from HCW (HCW20, HCW18, HCW36) as well as inpatients (p2 and p3) per 50μl of PBS. The mixtures above were incubated for 30 min at room temperature, then a suspension of 0.5% turkey red blood cells (RBC, Lampire Biological) was added, and plates were further incubated for 1 hour at 4°C. A/Michigan/45/2015 (matching with seasonal vaccine) was used as the reference strain to assess antigenic drift in the clinical viral isolates. Likewise, immune sera from ferrets exposed to A/Michigan/45/2015 were used for this purpose. Hemagglutination titers were calculated as the reciprocal of the last serum dilution at which the antibodies present in the serum inhibited agglutination of the viruses with the red blood cells.

## Data Availability

The data referred to in the manuscript will be made available upon request

## Acknowledgments

We thank the numerous team members of the Clinical Microbiology laboratories at the Mount Sinai Health System and the Department of Microbiology for expert support and their willingness to go the extra mile. We thank Ms. M.C. Bermudez and the team of the Personalized Virology Initiative for expert processing of human samples. We also extend our gratitude to Dr. A. Garcia-Sastre and Dr. P. Palese for guidance and many thought provoking discussions. We are indebted to Dr. C. Cordon-Cardo for continued support of the Pathogen Surveillance Program.

We would like to thank the staff, the Nursing leadership, including Christine Mahoney and Maria Latrace, the Residency Program Leadership including Daniel Steinberg, the Hospital Leadership including Dr. J. Boal at Mount Sinai Beth Israel, and the New York State Department of Health, especially Dr. K. Southwick and Rafael Fernandez, for their extraordinary help and support during the outbreak.

We gratefully acknowledge the authors, originating and submitting laboratories of sequences from GISAID’s EpiFlu (www.gisaid.org) that were used as background for our phylogenetic inferences. The list of authors and submitting laboratories is shown in **Table S1**.

This study was supported in parts by the National Institute of Allergy and Infectious Disease (NIAID) Centers of Excellence for Influenza Research and Surveillance (CEIRS) contract HHSN272201400008C, the Office of Research Infrastructure of the National Institutes of Health (S10OD018522 and S10OD026880) and institutional seed funds.

This manuscript was edited by Life Science Editors.

## Authors contributions

J.W, J.E, B.K., B.B., M.M, and I.M. were involved in the outbreak investigation. J.W, J.E, M.G., and E.M.S. provided clinical evaluations. J.W, J.E, M.G., and E.M.S. collected and verified clinical data. E.H., J.T, T.L., F.C., L.P., M.M.H., M.G., E.M.S., V.S., and H.B. performed clinical sample accessioning. E.H., M.M.H., and H.A. performed RNA extraction and viral subtyping. J.M.C, and V.S. selected and grew viral isolates. R.A.A., and V.S. provided banked serum samples. J.M.C performed HAI assays. A.S.GR., Z.K., and L.M. performed NGS experiments. A.S.GR., R.S., and M.S. provided NGS services. A.S.GR., D.K., T.R.P., and H.B. performed genome assembly and/or comparative genome analyses. J.E, T.R.P., E.M.S., and H.B. performed mining of electronic medical records. J.W, J.E, A.S.GR., M.M.H., F.K., E.M.S., V.S., and H.B. analyzed, interpreted, and/or discussed data. A.S.GR., E.M.S., V.S., and H.B. wrote the manuscript. E.M.S., V.S., and H.B. conceived the study. E.M.S., V.S., and H.B. supervised the study. E.M.S., V.S., and H.B. raised financial support

